# Accuracy and Acceptability of Wearable Motion Tracking Smartwatches for Inpatient Monitoring

**DOI:** 10.1101/2020.07.24.20160663

**Authors:** Chaiyawan Auepanwiriyakul, Sigourney Waibel, Joanna Songa, Paul Bentley, A. Aldo A. Faisal

## Abstract

Inertial Measurement Units (IMUs) within an everyday consumer smartwatch offer a convenient and low-cost method to monitor the natural behaviour of hospital patients. However, their accuracy at quantifying limb motion, and clinical acceptability, have not yet been demonstrated. To this end we conducted a two-stage study: First, we compared the inertial accuracy of wrist-worn IMUs, both research-grade (Xsens MTw Awinda, and Axivity AX3) and consumer-grade (Apple Watch Series 3 and 5), relative to gold-standard optical motion tracking (OptiTrack). Given the moderate to strong performance of the consumer-grade sensors we then evaluated this sensor and surveyed the experiences and attitudes of hospital patients (N=44) and staff (N=15) following a clinical test in which patients wore smartwatches for 1.5– 24 hours in the second study. Results indicate that for acceleration, Xsens is more accurate than the Apple smartwatches and Axivity AX3 (RMSE 0.17±0.01 g; R^2^ 0.88±0.01; RMSE 0.22±0.01 g; R^2^ 0.64±0.01; RMSE 0.42±0.01 g; R^2^ 0.43±0.01, respectively). However, for angular velocity, the smartwatches are marginally more accurate than Xsens (RMSE 1.28±0.01 rad/s; R^2^ 0.85±0.00; RMSE 1.37±0.01 rad/s; R^2^ 0.82± SE 0.01, respectively). Surveys indicated that in-patients and healthcare professionals strongly agreed that wearable motion sensors are easy to use, comfortable, unobtrusive, suitable for long term use, and do not cause anxiety or limit daily activities. Our results suggest that smartwatches achieved moderate to strong levels of accuracy compared to a gold-standard reference and are likely to be accepted as a pervasive measure of motion/behaviour within hospitals.

## 1. Introduction

Wearable movement sensors have the potential to transform how we measure clinical status and wellbeing in everyday healthcare. Tracking patient movements can help characterise, quantify, and monitor physical disability; highlight deteriorations; and signal treatment response [1]. Remote patient assessment may also allow for more cost-effective monitoring and offer advantages in contexts where direct contact is restricted e.g. due to Covid-19 associated isolation. Currently, behavioural assessments in clinical settings are characterized by intermittent, time-consuming human observations, using inconsistent subjective descriptions [1]. With an ageing population and increasing health system costs, there is a growing interest in seeking low-cost, automated methods for observing and quantifying patient behaviours [2-7]. Presently, the two leading wearable sensors offered for automated motion tracking are 1) camera-based optical tracking systems and 2) body-worn Inertial Measurement Units (IMUs), consisting of a triaxial accelerometer, a triaxial gyroscope, and, frequently, a magnetometer, that record linear accelerations and angular velocities in a three-dimensional (3D) Cartesian space.

Body-worn IMUs hold several advantages over optical systems for behaviour tracking ‘in the wild’ (i.e. free-living conditions). While optical systems are considered the gold-standard for high-accuracy movement tracking in controlled laboratory environments, the restriction on cameras’ field of views, obscuration of reflective markers, and lighting confounds [4] render these systems impractical for clinical use. Furthermore, optical equipment is expensive, cumbersome, complex to calibrate and operate, and has limited usage duration [4]. In contrast, IMU sensors offer a low cost, highly portable, robust, and inconspicuous alternatives that are better suited to measure daily life activities in unconstrained environments such as hospitals and care homes [8-10]. A diverse range of wearable IMUs is commercially available, which can be broadly grouped into consumer-grade products such as wrist-worn fitness trackers or smartwatches and research-grade IMU sensors for research or clinical purposes [11]. Consequently, widely adopted commercial products with networking functionalities are increasingly being applied for motion tracking applications with advantages of being ubiquitous, relatively low-cost, robust, easily cleanable, and simple to self-apply and operate [11-13]. Both fitness bands and smartwatches fall within this wearable category. We focus here on smartwatches as they are more easily programmable and facilitate distribution and update of custom software through App stores, thus making them attractive as a platform for wearable research and development. Smartwatches are already increasingly employed for health monitoring purposes, and so there is a growing need to assess their measurement precision against gold-standard references.

Presently, the use of consumer smartwatches in health applications is limited by the unknown data quality of their IMU data and their evaluation in a research or clinical setting. Previous work [13-20] focused on validating built-in heart rate, energy expenditure, and step count measurements relative against ground-truth measurements of electrocardiography, indirect calorimetry, and observed step counts. For instance, Wallen and colleagues [14] found that the smartwatches, Apple Watch (Apple Inc., Cupertino, CA), Fitbit Charge (Fitbit Inc., San Francisco, CA), Samsung Gear S (Samsung, Seoul, South Korea) and Mio Alpha (MioLabs Inc., Santa Clara, CA), underestimated outcome measurements such as step count in terms of average error range between 4% to 7% (Apple Watch error = −4.82%, Fitbit Charge HR error = −5.56%, and Samsung Gear S error = −7.31% (relative errors computed from the raw data provided in the paper).

In evaluating accuracy and precision of smartwatch IMUs, however, both absolute errors (i.e. how much is the sensor differ from the ground-truth value) and correlations (i.e. how well does the sensor track the dynamic changes of ground-truth values) need to be measured. Apple Watch (r = 0.70), Fitbit Charge HR (r = 0.67) and Samsung Gear S (r = 0.88) were shown to correlate reasonably well with ground-truth step count [14]. Note, out of the 3 smartwatches, the one with the highest average error also shows the best performance in tracking step count dynamically, so signal quality rankings for the same watch differs across accuracy and precision measures. Moreover, across sensing modalities that were not related to kinematics, the same set of watches performed different in how well they captured heart rate and energy expenditure, and no single smartwatch was the best in overall assessed modalities.

Validating smartwatch derived measures for clinical or scientific use is complicated as most measured outcomes recorded from consumer wearables e.g. built-in energy expenditure and step counts are derived from undisclosed, proprietary algorithms with unknown modelling assumptions that have not gone through medical certification processes. Assuming that different generations of the same smartwatch models are not significantly different from each other across studies, it suggests that experimental protocols play a considerable impact in assessing measurement quality. Therefore, the assessment and direct comparison of devices require a defined and reproducible description of the assessment process. Another study [19], compared fitness armbands and smartwatches to the gold-standard measurements, found that even average step count errors varied widely between wearable devices, such as Nike+ FuelBand (Nike Inc., Beaverton, Oregan) (error = 18.0%), Jawbone UP (Jawbone, San Francisco, CA) (error = 1.0%), Fitbit Flex (Fitbit Inc., San Francisco, CA) (error = 5.7%), Fitbit Zip (Fitbit Inc., San Francisco, CA) (error = 0.3%), Yamax Digiwalker SW-200 (Yamasa Tokei Keiki Co. Ltd, Tokyo, Japan) (error = 1.2%), Misfit Shine (Misfit Wearables Inc., San Francisco, CA) (error = 0.2%), and Withings Pulse (Withings, Issy-les-Moulineaux, France) (error = 0.5%), showing that even wearable sensors developed specifically for tracking fitness activities such as walking and running activity made some errors, although it is unclear if a more generous counting of steps may be of fitness or commercial interest.

The lingering uncertainty in the quality and usability of the underlying sensor signal quality motivated our work here, by first evaluating the raw IMU sensor data quality of smartwatches, and then second, trial the feasibility of large-scale deployment in a clinical care setting through the patient (PPI) and healthcare worker involvement. Clinical and care wearable applications and analysis rely upon quantified, regulatory acceptable measures of accuracy of the fundamental signal (linear acceleration for accelerometers and angular velocity for gyroscopes) that must be compared to gold standards derived from marker-based optical motion tracking. The quality in these fundamentals signal allow us to assess how well they can in principle track measures, such as body kinematics, but also more indirectly inferred measures, often clinically outcome measures and primary endpoints of clinical trials (such as step counts). To date, there has been no independent direct comparison between common consumer smartwatches and research-grade IMUs inertial accuracy relative to ground truth optical motion tracking. This is in part due to consumer smartwatch closed-system barriers to raw IMU data extraction, which we overcome through developing customised software. We also developed an easily reproducible measurement protocol to directly assess and compare smartwatches in naturalistic movement tasks, performed by the same human on all compared devices at the same time. Additionally, a separate issue for the clinical feasibility of smartwatches in care is the feasibility of their deployment is their practicality and acceptability in everyday use by both patients and healthcare workers. Research to date on patient and healthcare staff attitudes towards the continuous wearing of IMU sensors is scarce. While some studies report user-perceptions (e.g. user-friendliness and satisfaction) of smartwatch and fitness devices [13,21-25], these often focus on community settings, chronic disease, young/middle-age subjects, and healthy participants and as such are not as relevant for typical in-patient populations.

We focussed among the consumer-grade smartwatches on a single smartwatch make, so we could evaluate the technology within a large, parallel deployment of units in the care setting while remaining within a reasonable budget. We used market share as a guide for deciding which smartwatch to evaluate: We chose the Apple Watch (47.9% market share), with the second most popular device, Samsung Gear, holding only 13.4% of the global market share (in terms of shipment units) in the first quarter of 2020 [26].

## 2. Materials and Methods

### 2.1. Sensor Signal Quality Study

#### 2.1.1. Population

We recruited a sample of healthy volunteers (n=12) from Imperial College London to take part in a sensor accuracy assessment study. All participants agreed to take part with no withdrawals. All participants gave informed consent to participate in the study, and the study was ethically approved by Imperial College London University Science, Engineering and Technology Research Ethics Committee (ICREC).

#### 2.1.2. Data Collection

To record and extract inertial data from the Apple Watches (Series 3 and 5), we developed a piece of software, a WatchOS App, to collect real-time triaxial acceleration (±8 g for Series 3 and ±16 g for Series 5) and triaxial angular velocity data (±1000 degree/s for Series 3 and ±2000 degree/s for Series 5) at 100Hz. The watch stored data to an onboard memory and offloaded the data to a custom-configured base station wireless access point and laptop. The Xsens MTw Awinda unit recorded packet-stamped triaxial acceleration (±16 g), triaxial angular velocity (±2000 degree/s), and triaxial magnetic fields (±1.9 Gauss) data at 100Hz. The Xsens sensors wirelessly transmitted data in real-time to a base station and laptop which recorded the data within the MT Manager software. The Axivity AX3 unit recorded triaxial acceleration data at 100Hz with a configurable range of ±2/4/8/16 g (±16 g was selected for this study). The Axivity sensors stored data to an onboard memory and offloaded the data upon attachment to a laptop via the installed AX3 OMGUI software. For the ground-truth optical motion tracking, we calibrated 4 ceiling-mounted OptiTrack cameras according to the manufacturer’s specifications and created a rigid body model using four reflective markers attached to the single marker pad (see photos in Fig. 1.a). The system recorded absolute x-axis position and triaxial rotation of the marker within a 2 cubic meters area at 240Hz and wirelessly recorded data in real-time within a laptop installed Motiv Software.

**Figure 1.**
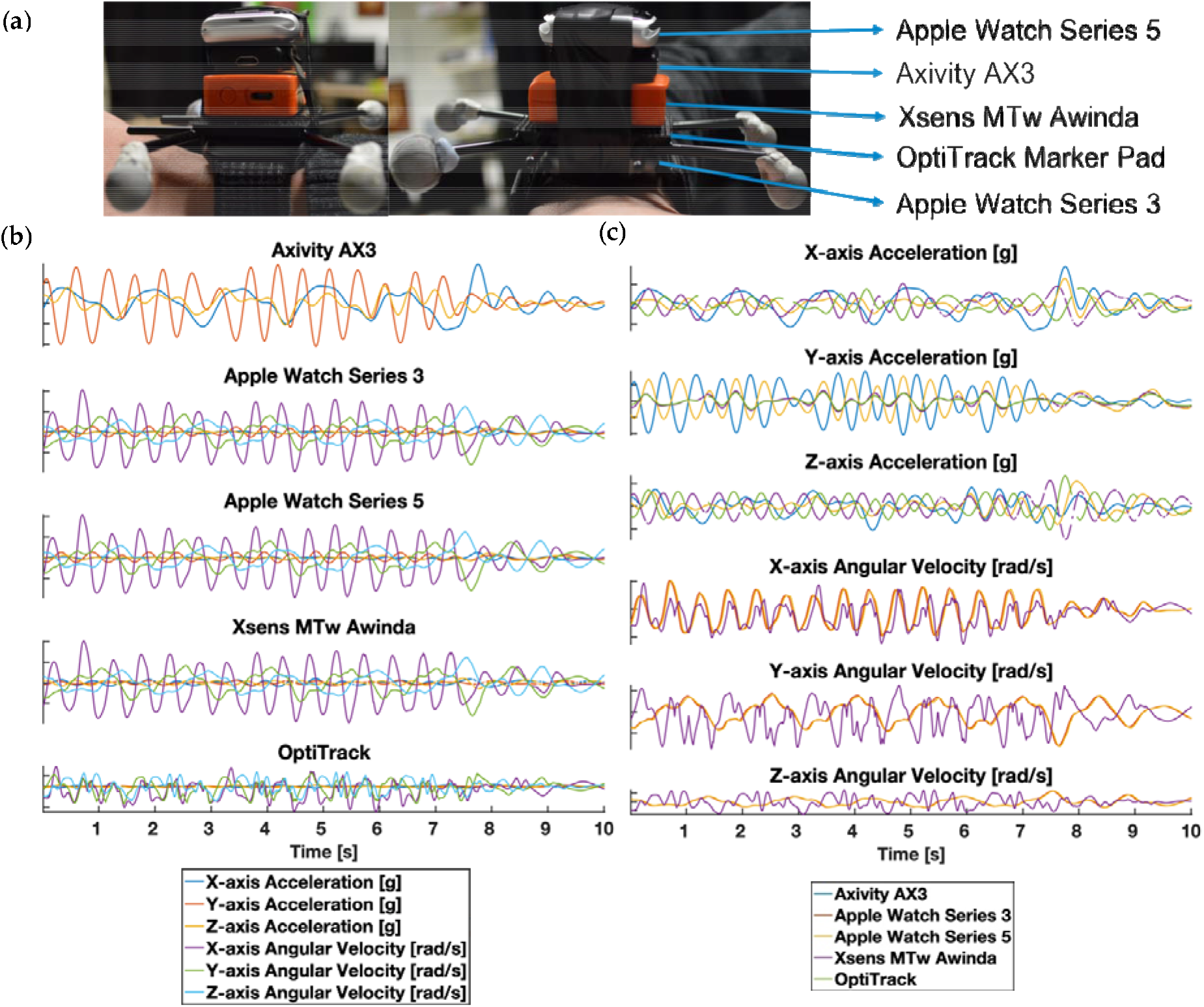
**(a)** depicts the participant wearing the sensor stack attached to the right wrist with the adjustable wrist strap. From top to bottom of the sensor stack, we mounted Apple Watches Series 5, Axivity AX3, Xsens MTw unit, OptiTrack retro-reflective marker pad with 4 retro-reflector unit (grey spheres), and Apple Watch Series 3 to one another vertically, aligning the centre of gravity, using Velcro sticky pads. (b) depicts 10-seconds sensors reading for each of the motion sensors. (c) shows 10-seconds sensors reading for each of the recording dimensions.

We asked each participant to perform a predefined sequence of upper body movements in a 6-minute controlled exercise while data were simultaneously recorded from the sensor and marker stack illustrated in Figure 1. We chose the movement tasks outlined in Table A.1 because they spanned the full range of natural joint angles at shoulder and elbow during a complex 2-joint movement typical for natural activities e.g. reaching for, passing, and picking up an object. Each participant trial consisted of a sequence of 4 distinct movements in time with a 120 BPM metronome. We constrained movement tasks within the OptiTrack cameras’ field of views.

#### 2.1.3. Data Processing

Following the completion of the movement protocol, we collected and analysed sensor inertial data in MATLAB® (MathWorks, Inc., Natick, MA, USA). All sensor data was linearly resampled to a constant sampling rate of 100Hz. We manually inspected the OptiTrack data within Motiv, corrected mislabelled markers, and reconstructed the rigid body data using the newly corrected makers. We derived OptiTrack triaxial accelerations and angular velocities from the positional and rotational data. We unrolled Xsens packet stamps and replaced missing packet rows with a Not-a-Number (NaN) row vector and calculated Xsens timestamps using the unrolled packet stamps.

A 0.25 to 2.5 Hz 6^th^-order Butterworth bandpass filter decontaminates signals data from integration drifts and differentiation noise. We determined the cut-off frequencies by plotting the power spectrums of acceleration (a) and position (b) signals of the sensors, which demonstrated that acceleration concentrate around 2 Hz and position around 0.5 and 1 Hz (Figure 2). This frequency is consistent with our movement tasks which were restricted to a lower bound of 0.5 Hz or an upper bound of 2 Hz. Additionally, we used Hampel filters to identify and interpolate outlying spikes in the OptiTrack data caused by differentiating missing frames and incorrectly inverted rigid body orientation resulting from marker obscuration.

**Figure 2.**
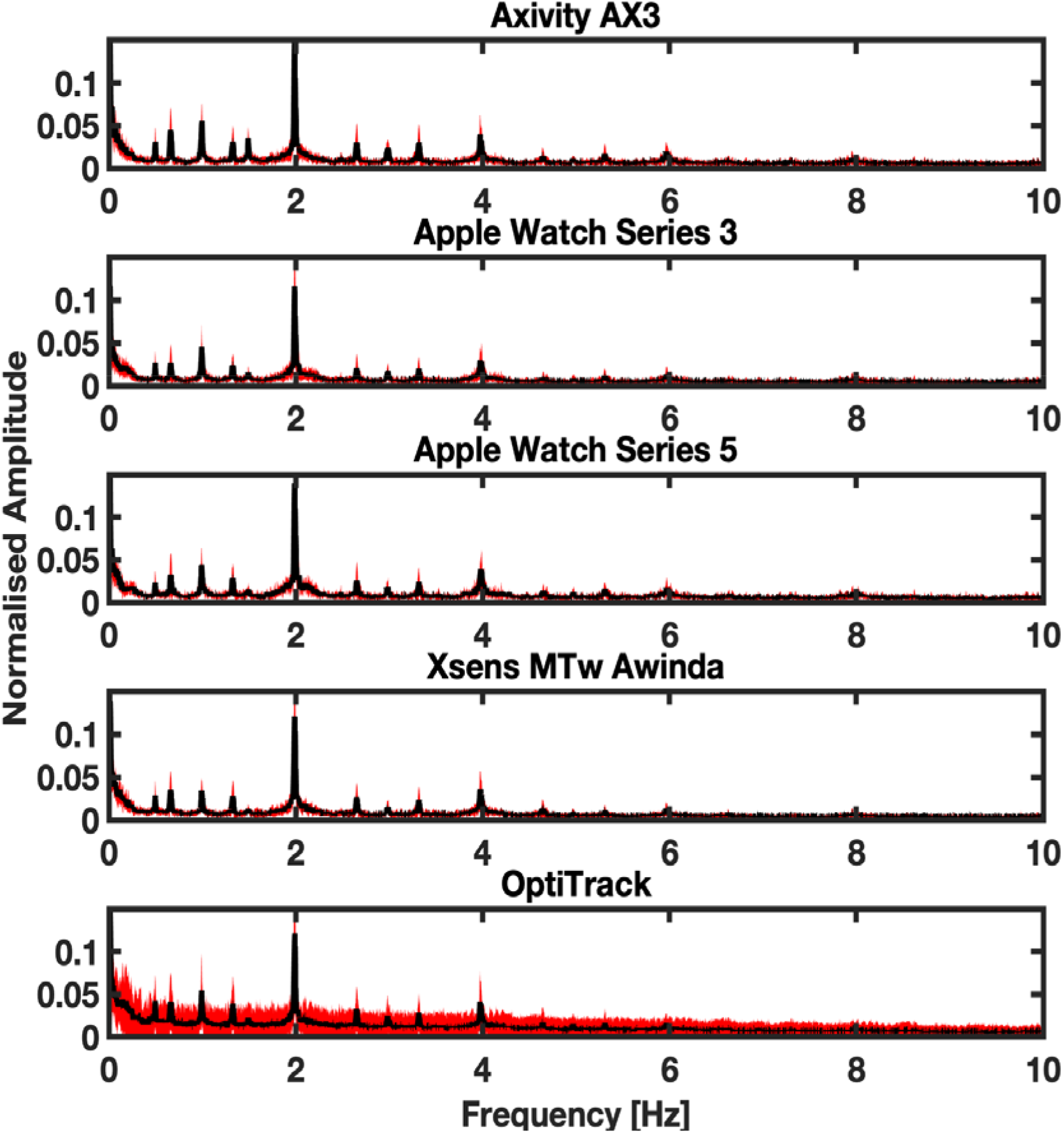
Depicts the power spectral density for the linear acceleration signal for each aggregated across all validation study participants. Shaded areas indicate 1 SD from the mean power, each row represents data from a different device. All evaluated device data and optical motion tracking were collected simultaneously.

**Figure 3.**
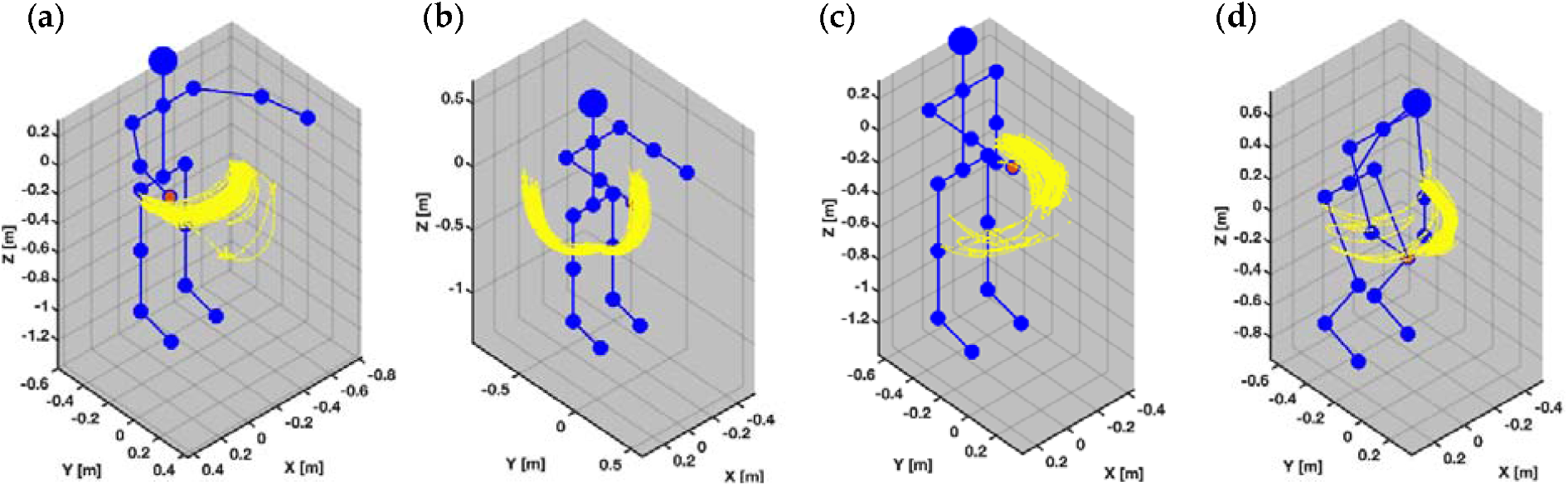
Depicts in blue the action component order for the participant performing the 4 movement tasks during the sensor validation protocol. Illustrates in yellow the OptiTrack reflective marker movement path associated with each movement task. From top to bottom, the 4 movement tasks are: (a) Horizontal Arm Movement, (b) Vertical Arm Movement, (c) Rotational Arm Movement, and (d) Composite Cross-Body Movement.

We visually segmented sensor recordings using an identifiable movement event (five handclaps) at the start and end of each participant recording. To align signals spatially, we converted acceleration data to a unit of earth gravity (g), using a constant of 9.80665 m/s^2^, and angular velocity data to a unit of radians per second (rad/s). We used cross-correlation to calculate any lag between signals in each unique device pair to align signals temporally at the common starting point. To verify alignment, we applied cross-correlation to each signal pair. We then converted aligned signals were into a 1D vector using a Euclidean norm function and removed NaN rows from each sensor vector and its corresponding sensor pair.

#### 2.1.4. Analysis

We compared vectors for each inertial sensor against every other inertial sensor and OptiTrack using Root-Mean-Square-Error (RMSE) and R-Squared (R^2^) metrics for acceleration and angular velocity. Axivity AX3 does not contain a gyroscope and so we omitted angular velocity comparisons for Axivity. We reported the results between each sensor pair in the format of Mean of Metrics across all trials ± Standard Error of Metrics across all trials. To assess whether the stacking of the sensors and markers introduced any recording error due to varied distance between the wrist and the sensor, we plotted the difference between acceleration and angular velocity from an Apple Watch (Series 3) stacked at the bottom and an Apple Watch (Series 5) stacked at the top of the sensor and marker stack. We interpreted the strength of the R^2^ between sensors using the following descriptive categories: weak (R^2^ = <0.5), moderate (R^2 =^0.5–0.7) and strong (R^2^≥0.7) agreement[14].

### 2.2. Sensor Acceptability Study

#### 2.2.1. Population

We recruited a consecutive convenience sample of 44 in-patients and 15 healthcare professionals to complete patient and healthcare professional sensor acceptability questionnaires, respectively. We recruited patients who were enrolled in a study exploring relationships between free-living wearable motion sensor data and observed behaviour and clinical scores (data to be published separately). Inclusion criteria were: 1) admission to Stroke or Neurology Wards at Charing Cross NHS Hospital, London; 2) > 18 years old; 3) absence of significant cognitive impairment; 4) Glasgow Coma Score >13; and 5) ability to understand English; 6) and capacity to consent. Exclusion criteria were: 1) inclusion within another interfering trial; and 2) contagious skin infections or other rash interfering with sensor placement. The study screened 71 potentially eligible patients, of which the following were subsequent reasons for further exclusion: swollen ankles (n=2); same-day discharge (n=5); and patient refusal to take part because they were not interested (n=7), or felt fatigued or unwell (n=8). 49 patients completed the study protocol, of which 5 were excluded due to: a clinical need to remove watches early to undertake magnetic resonance imaging (MRI) (n=2); and hospital discharge or transfer (n=3). All participants gave informed consent to participate in the study and the study was ethically approved by the UK’s Health Research Authority IRAS Project ID: 78462.

#### 2.2.2. Data Collection

All healthcare professional and patient interviews took place in the ward by the patient bedside. The clinical researchers explained the purpose of the study both verbally and via written information sheets. Questionnaires lasted 10-minutes and two clinical researchers recorded the participants’ answers.

All in-patients wore 4 consumer smartwatches (Apple Watch Series 3) on their wrists and ankles (i.e. 1 per limb) while they performed their usual everyday activities on the hospital ward. We chose this arrangement to capture asymmetrical patterns of, upper, and lower limb weakness (that are typical for stroke); and to support recognition of both different locomotion (e.g. walking, standing and sitting) and different daily activities entailing manual interactions (e.g. drinking and eating). To assess sensor wear-time acceptability over day-long continuous recording periods, we asked all participants to wear the sensors for a full working day and a random subset of the total (n=11) to continue to wear the sensors overnight. Subject wear times varied (1.5-24 hours), as watches were removed for patient showers, medical scans, and upon patient hospital discharge or transfers. At the end of the recording protocol, we asked participants to complete a questionnaire to collate their views regarding wearable technology. We derived and adapted [24,27,28] and agreed upon the final study questions via discussions between the clinical researchers, a Consultant Neurologist, a Stroke Physician and two patients. Watches were attached using a soft, breathable nylon replacement sport strap with adjustable fastener. During the protocol, we locked watch user-functionalities, blanked out and covered the watch screen with a plastic sleeve preventing user interaction. After sensor recordings, in-patients answered 10 close-ended questions outlined in Table 1.

**Table 1.** Healthcare Professional and in-patient Questionnaire

| **In-patient Closed Questions** |  |
| --- | --- |
| • icQ1. | The device was easy to put on and take off? |
| • icQ2. | I would feel comfortable wearing the device even if it is visible to others? |
| • icQ3. | I feel I could do most of my normal activities (except those involving water) wearing the device? |
| • icQ4. | The device did not interfere with washing or going to the toilet? |
| • icQ5. | I would find it easy to learn to use the device? |
| • icQ6. | I did not experience any itchiness or skin irritations using the device? |
| • icQ7. | I did not experience any discomfort wearing the device? |
| • icQ8. | I did not feel anxious wearing the device? |
| • icQ9. | I would be willing to wear the device continuously for long term use? |
| • icQ10. | I did not find the appearance or design of the sensors obtrusive? |
| **Healthcare Professional Closed Questions** |  |
| • hcQ1. | The device was easy to put on and take off? |
| • hcQ2. | I would find it easy to learn to use the device? |
| • hcQ3. | Do you think that the increasing use of wearable tracking technology and Artificial Intelligence in healthcare is an opportunity? |
| • hcQ4. | Do you think that the increasing use of wearable tracking technology in Artificial Intelligence in healthcare is a danger? |
| • hcQ5. | If there were strong clinical evidence that the intervention would be equivalent or better than current neurological observations alone in a Neurology and Stroke setting, would you agree to use the new intervention in your own management of your patients? |
| **In-patient Open Questions** |  |
| • ioQ1. | What do you like about the device? |
| • ioQ2. | What sort of characteristics and functions do you expect from the device? |
| • ioQ3. | Is there anything you don't like about the device? |
| **Healthcare Professional Open Questions** |  |
| • hoQ1. | What do you like about the device? |
| • hoQ2. | What sort of characteristics and functions do you expect from the device? |
| • hoQ3. | Is there anything you don't like about the device? |
| • hoQ4. | What do you think are the benefits and risks you perceive when using these new technologies? |

Q =question, ic =in-patient closed; hc =healthcare professional closed, io =in-patient open; ho =healthcare professional open

We provided healthcare professionals with the intended functionality of the sensors for monitoring patient movement in the hospital and showed healthcare professionals how to operate the devices. Healthcare professionals then answered 5 closed-ended questions outlined in Table 1. Thereafter, we asked both in-patients and healthcare professionals open-ended questions described in Table 1.

Rating scales and descriptive category groupings were as follows:

- icQ1-10 used a 1 to 7 rating scale: 1 to 2 (strongly disagree); 3 to 4 (somewhat agree); and 5 to 7 (strongly agree).
- hcQ1-2 used a 1 to 7 rating scale: 1 to 2 (strongly disagree); 3 to 4 (somewhat agree); and 5 to 7 (strongly agree).
- hcQ3 used a 1 to 10 rating scale: 0 to 5 (no opportunity); and 6 to 10 (great opportunity).
- hcQ4 used a 1 to 10 rating scale: 0 to 5 (no danger/safe); and 6 to 10 (danger).
- hcQ5 was collected with a −3 to +3 rating scale: −3 (would not use the intervention); −2 to 0 (would only use the intervention if controlled by a human caregiver); and 1 to 3 (would use the intervention and it could replace some interventions currently implemented by human caregivers).

### 2.2.3. Analysis

We aggregated in-patient and healthcare professional responses for close-ended questions into the broader rating scale descriptive categories (e.g. strongly disagree, somewhat agree, strongly agree for the 1-7 rating scale) and calculated 1) the percentage of responses for each descriptive category for in-patients; and 2 the frequency of responses for each descriptive category for healthcare professionals separately. We assessed all open-ended responses by a thematic analysis which aimed to describe concepts extracted from the participant responses. Literal comments were 1) recorded by the two clinical researchers 2) compared and grouped based on similarity and creation and 3) subsequently merged into core agreed themes via consensus between the researchers.

## 3. Results

Both the sensor signal quality comparison and the sensor acceptability study we explore here are based on the Apple Watch (Series 3 and 5), which we chose based on its market share as in 2019/2020 (47.9% market share) as the single leading consumer device, as a second most used device in the market had only 13.4% market share [26]. We compared the inertial accuracy of the consumer smartwatch IMUs (from Apple Watch Series 3 and 5) against two well-known research- and clinical-grade IMU sensors: 1. Xsens MTw Awinda (Xsens Technology B.V., Enschede, The Netherlands) with many published applications and validation studies in biomechanics (e.g. [29-35]) and 2. Axivity AX3 (Axivity Ltd, Newcastle Upon Tyne, UK)[36,37] with many published biomedical research applications including its deployment in the UK Biobank cohort with over 3,500 devices used by 100,000 participants (e.g.[38]).

We determined how all three IMUs compared to a gold-standard for human movement assessment in the form of optical motion tracking (OptiTrack, Natural Point Inc., Corvallis, OR) [39-41]. The findings from the first study led to the selection of the smartwatch sensors for the second study in which we conducted a survey, assessing the experiences and attitudes of 44 in-patients and 15 healthcare professionals after a trial of continuous smartwatch use in hospital patients. Together these questions establish the scientific and practical validity of wearable inertial sensors for movement tracking in clinical applications, particularly within hospitals.

### 3.1. Sensor Signal Quality Study Results

We compared the inertial accuracy of both research-grade and consumer-grade IMUs relative to gold-standard optical motion tracking. The RMSEs for acceleration against OptiTrack ranged from 0.15 to 0.42 m/s^2^ and RMSEs for angular velocity ranged from 1.28 to 1.40 rad/s. When comparing to OptiTrack acceleration, R^2^ agreement was stronger for Xsens than for Axivity, the latter of which had weak R^2^ agreement. Apple Watches only demonstrated moderate R^2^ agreement with OptiTrack acceleration. When comparing to OptiTrack angular velocity, R^2^ agreement was similarly strong for Xsens and Apple Watch (Series 3 and 5). When comparing to Xsens (research inertial sensor reference) acceleration, Apple watch demonstrated stronger R^2^ agreement than Axivity. When comparing to Xsens (research inertial sensor reference) angular velocity, Apple watch (Series 3 and 5) had strong R^2^ agreement. Apple Watch Series 3 and Apple Watch Series 5 had a strong agreement with each other for acceleration and angular velocity: The actual R2 and RMSE values for acceleration comparisons (Figure 4a & b) and RMSE value for angular velocity comparisons (Figure 4c & d) between Apple Watch Series 3 and Series 5 (which in the graphic are rounded to two digits) are 0.9997±0.00001, 0.0049±0.00016, and 0.9999±0.00000 respectively, these are not visible in the figures due to rounding. The actual RMSE value for angular velocity comparisons between Apple Watch Series 3 and Xsens MTw Awinda is 0.9955±0.00030.

**Figure 4.**
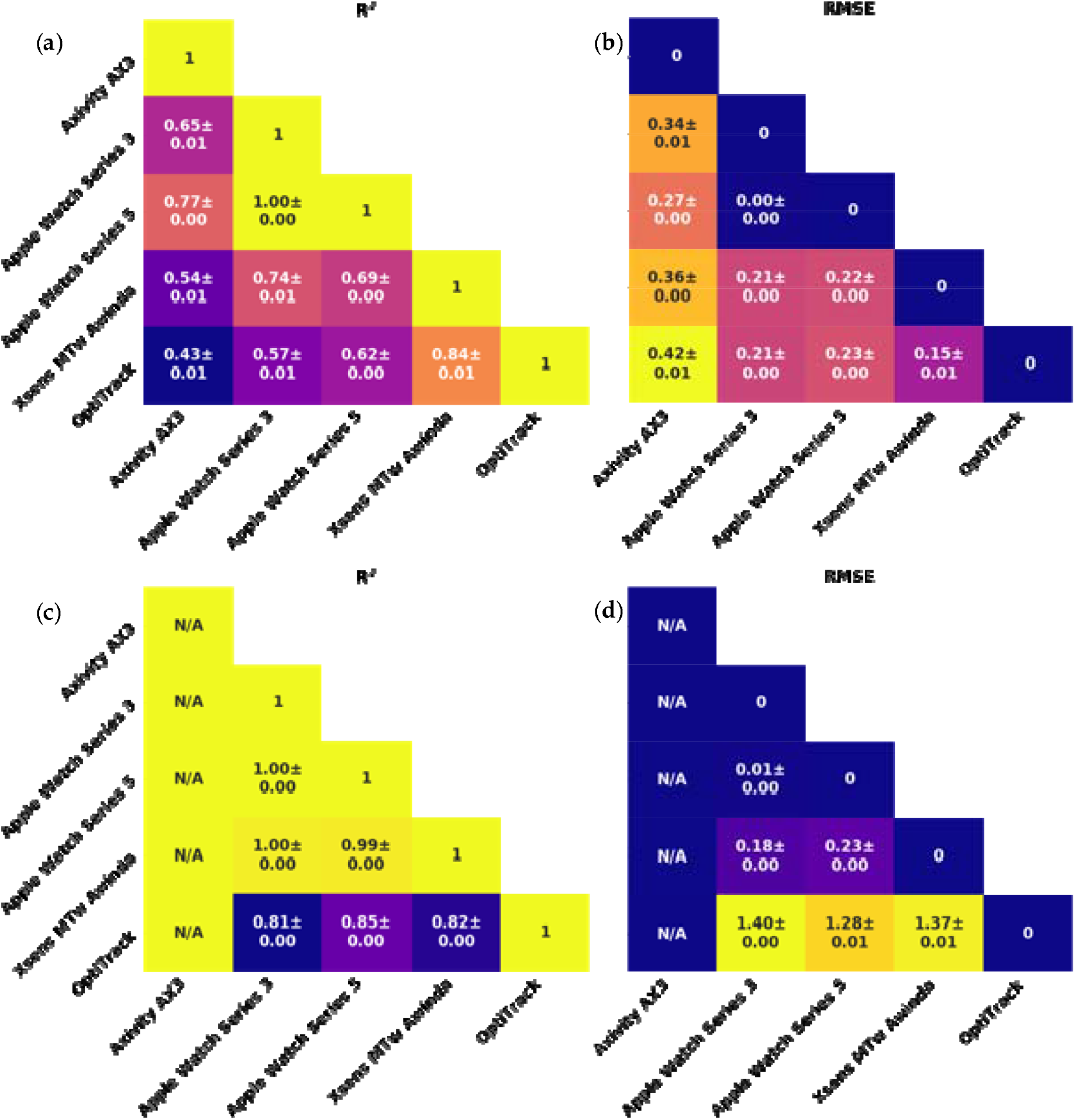
Triangle diagrams of the sensor validation study aggregated by pairwise signal measures (R^2^ for dynamic tracking of signals and RMSE metrics for offsets) between the selected motion sensors (Apple Watch Series 3 and 5, Xsens, Axivity, and Optitrack). Data is organised in (a) linear accelerometer R^2^, (b) linear accelerometer RMSE, (c) angular velocity R^2^, and (d) angular velocity RMSE. Displayed R^2^ and RMSE values in the figure are rounded. See main text for details.

**Figure 5.**
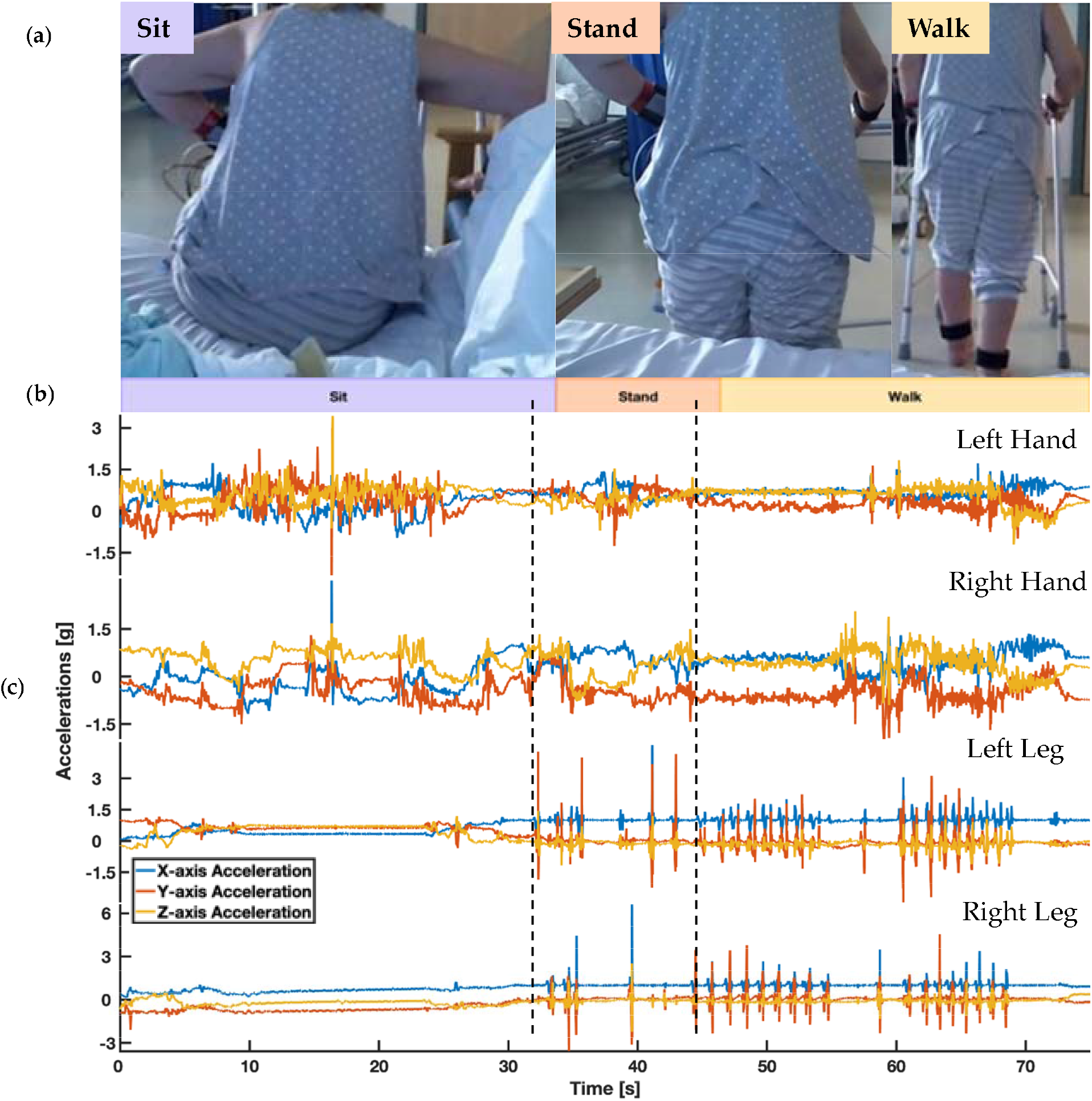
Depicts the participant’s behaviour while wearing the 4 wrist and ankle-worn sensors on the hospital ward and the associated sensors’ triaxial acceleration readings. (a) depicts the ground-truth video recording of the subject. (b) describes the associated behaviour labels. (c) shows the associated 3D linear acceleration signals of the 4 sensors.

### 3.2. Sensor Acceptability Study Results

A total of 44 patients (50% female; average age: 64 years; interquartile age range: 24-92 years) completed the acceptability questionnaire. A further 15 healthcare professionals (66% female; doctors =5, nurses = 4, therapists = 3, and healthcare assistants = 3) working directly with these patients were also recruited. Further details of in-patient and healthcare characteristics are outlined in Table A3 and Table A4 respectively.

The patient survey responses showed that in-patients strongly agreed with all 10 closed-ended questions (icQ1-10), suggesting that sensors were easy to operate and learn to use, comfortable, did not limit daily activities, did not cause anxiety, and unobtrusive in appearance as seen in Table 2. As illustrated below in Table 2, the survey of healthcare professional showed high levels of agreement with statements that the system was easy to operate and learn to use (hcQ1-2) and presented no danger (hcQ4). Healthcare professionals were more split in their views regarding the opportunity of wearable tracking sensors and AI in healthcare delivery (hcQ3) and whether the technology could be used without the control of a human caregiver (hcQ5). Difference in opinions existed across the varied healthcare professional specialties. In particular, given strong evidence that an intervention was better or equivalent to current observations, some therapists (n=2), nurses (n=3), and healthcare assistants (n=1) still viewed human control as important, whereas all doctors (n=5) viewed human control as unnecessary. Additionally, all therapists (n=3) viewed the increasing use of wearable motion sensors and artificial intelligence technologies as an opportunity for healthcare applications, whereas some doctors (n=3), nurses (n=2), and healthcare assistants (n=1) thought it presented no opportunity. A significant proportion of in-patients reported that they felt neutral towards the sensors or had nothing in particular to comment when asked open questions about system likes (n=18), dislikes (n=34), and expected functions and characteristics (n=31) (ioQ1-3). The comments from in-patients who did provide detailed responses were grouped into various themes (5 likes, 5 dislikes, 6 expected characteristics and functions) outlined in Table 3.

**Table 2.** Closed-ended Question Results for In-patients and Healthcare Professionals

| In-patient Questionnaires | Strongly Disagree | Somewhat Agree | Strongly Agree |
| --- | --- | --- | --- |
| • icQ1. | 2 | 2 | 39 |
| • icQ2. | 0 | 5 | 38 |
| • icQ3. | 0 | 2 | 41 |
| • icQ4. | 4 | 2 | 38 |
| • icQ5. | 2 | 4 | 37 |
| • icQ6. | 1 | 1 | 42 |
| • icQ7. | 2 | 2 | 40 |
| • icQ8. | 2 | 0 | 42 |
| • icQ9. | 3 | 5 | 36 |
| • icQ10. | 1 | 3 | 39 |
| **Healthcare Professional Questionnaires** | **Strongly Disagree** | **Somewhat Agree** | **Strongly Agree** |
| • hcQ1. | 2 | 0 | 13 |
| • hcQ2. | 2 | 0 | 13 |
|  | **No Opportunity** | **Great Opportunity** |  |
| • hcQ3. | 6 | 9 |  |
|  | **Dangerous** | **Safe** |  |
| • hcQ4. | 0 | 15 |  |
|  | **Would not use** | **Would only use if human-controlled** | **Would use and replace** |
| • hcQ5. | 0 | 6 | 9 |

Table 2 describes 1) the percentage of responses for each descriptive category for in-patients (n=44); 2) the frequency of responses for each descriptive category for healthcare professionals (n=15). The percentages may not add up to 100% due to the rounding error. icQ1-10 and hcQ1-2 used a 1 to 7 rating scale: 1 to 2 (strongly disagree); 3 to 4 (somewhat agree); and 5 to 7 (strongly agree). hcQ3 used a 1 to 10 rating scale: 0 to 5 (no opportunity); and 6 to 10 (great opportunity). hcQ4 used a 1 to 10 rating scale: 0 to 5 (no danger); and 6 to 10 (danger). hcQ5 was collected with a -3 to +3 rating scale: -3 (would not use the intervention); -2 to 0 (would only use the intervention if controlled by a human caregiver); and 1 to 3 (would use the intervention and it could replace some interventions currently implemented by human caregivers).

**Table 3.** Themes of perceived likes, dislikes, expected functions and characteristics of technology from in-patient survey (ioQ1-3)

| Themes | Details and example quotes |
| --- | --- |
| **Likes** |  |
| • Attractive appearance | 'Fine', 'good', 'stylish', 'beautiful', 'modern', design |
| • Unobtrusive | 'unobtrusive', 'neutral' or 'unaware' of the device. Sensor felt just like a 'normal watch', 'non-invasive' |
| • Simple design and ease of use | 'Simple design', 'easy to wear', 'simple to wear' |
| • Visibility to others | Not concerned about the visibility of sensors to others as 'appearance is fine' |
| • Promising healthcare applications | 'Helpful for research' and can improve healthcare |
| **Dislikes** |  |
| • Poor appearance | Would like a 'colour scheme' |
| • Discomfort | Skin 'irritation' from sensors |
| • Interference with medical equipment | Cumbersome to wear with other 'medical contraptions' |
| • Poor straps | Would like 'stretchy' and 'magnetic straps', straps 'hard to get on' |
| • Large size | 'too big' |
| **Expected characteristics and functions** |  |
| • Well-designed straps | 'Better looking' straps |
| • Attractive appearance | 'beautiful design', 'brighter' colour scheme |
| • Time and heart rate functionalities | 'heart rate' and 'time' functionalities |
| • Suitability for medical scans | 'Suitable' to wear when going for 'MRI scans' |
| • Promising healthcare applications | 'helpful for research', 'beneficial for other patients' |
| • Smaller size | 'smaller' size |

In response to the open-ended question (hoQ1-4), healthcare professionals (n=10) highlighted that the sensors would only cause discomfort to a selection of patients in certain situations (e.g. some cases of hemiparesis, swelling or long wear periods). All healthcare professionals viewed the system as not intrusive to healthcare professionals; and, similarly, the majority of healthcare professionals (n=12) also perceived that the sensors were not intrusive to patients. 6 healthcare professionals commented that the system may interfere with medical treatments, while 8 disagreed and thought it would not interrupt care needs. The open-ended comments from healthcare professionals were grouped into themes (7 benefits, 5 risks, 2 likes, 2 dislikes, 4 expected characteristics and functions) outlined in Table 4.

**Table 4.** Themes of perceived benefits, risks, likes, dislikes, expected functions and characteristic of technology from the healthcare professional survey (hoQ1-4)

| Themes | Details and example quotes |
| --- | --- |
| **Benefits** |  |
| Promising healthcare applications<br>Treatment personalisation<br>Patient engagement<br>Patient tracking<br>Unobtrusiveness<br>Ease of use<br>Convenience | 'Able to monitor movements to the development of new therapy'<br>'tailored therapy'<br>'way of engaging with patients in their own health'<br>'wearer could be tracked' to know 'where they are'<br>'gather information....in objective way & patients didn't seem inconvenienced'<br>Ease of use and quick set-up<br>'convenient in the modern days of medicine' |
| **Risks** |  |
| Data privacy risks<br>Sensor loss<br>Discomfort<br>Specificity<br>Compliance | 'ability for it to be shared with others that a patient did not consent to'<br>'it can be lost as it is easy to remove'<br>'not comfortable on skin and can contribute to skin wounds'<br>'risk of false-positive results'<br>use...' depends on patient compliance' |
| **Likes** |  |
| Ease of use<br>Promising healthcare applications<br>Lightweight<br>Unobtrusive<br>High portability | 'easy to wear and use'<br>Purpose and aim of system for health monitoring<br>Lightweight size<br>System is not obtrusive for patients or healthcare professionals<br>It's 'portable' so it is possible to 'monitor' whilst the patient is 'mobile' |
| **Dislikes** |  |
| Large size<br>Unsuitable for medical scans | 'too bulky'<br>frustrated by having to 'remove' the sensors to accommodate 'medical scans' |
| **Expected characteristics and functions** |  |
| Alarm systems<br>Integration vital measurements<br>On-screen instructions<br>Notifications | alerts to dangerous changes in 'symptoms' e.g. 'GCS scores' and area breaches within the ward<br>combine with other important clinical measurements e.g. 'temperature' and 'peripheral capillary oxygen saturation'.<br>'Instructions' on how to use device.<br>On-screen 'notifications' on device |

## 4. Discussion

Wearable inertial sensors are increasingly exploited for clinical purposes by providing low-cost, pervasive, high-resolution tracking of natural human behaviour. However, their validity assumes that they convey accurate motion information, while their clinical feasibility and adoption require a minimal level of user acceptance among patients and healthcare professionals. In this study, we tested these two assumptions by: 1) quantifying the accuracy of commonly used wearable inertial sensors relative to a gold-standard optical motion tracking instrument; and 2) surveying the attitudes of target healthcare professionals and in-patient populations following a trial period of continuous wearable inertial sensor use. As our study approached two original research questions, our results are not directly comparable to results of earlier literature as we addressed different research problems (raw sensor data quality) and employed standardised and consistent methods for collecting comparative data across devices. For example, we differed in our choice of outcome measure (we used straightforward inertial estimates rather than combined estimates (e.g.[29,32]) and type of sensor (we used individual sensors rather than full-body sensor suits(e.g.[29,32]). This is significant given that many motion tracking applications of clinical use, such as in-patient seizure detection, and sleep and posture classification [42] models depend upon using good raw accelerometer and gyroscope data.

### 4.1. Sensor Signal Quality Study

Relative to ground-truth optical motion tracking, the consumer smartwatches (i.e. the Apple Watch Series 3 and 5) and the research-grade IMU Xsens achieved cleaner linear acceleration signals and lower errors than Axivity (Figure 4). This is likely to be due to accelerometer and gyroscope fusion in the cases of Xsens and consumer smartwatches that enables superior isolation of gravity vectors from acceleration signals; whereas Axivity acts as a pure acceleration logger that relies on a low-pass filter to accomplish the [43-45]. We found that the consumer smartwatches and Xsens sensors had similarly high angular velocity accuracy when compared against ground truth (Figure 4). However, Xsens had stronger fidelity for recording accelerations (R^2^=0.88), perhaps due to the additional magnetometer and strap down integration (SDI) technology [46-49]. Accelerations and angular velocities were for all purposes identical between Apple Watch Series 3 and Series 5 (Figure 4), suggesting high intergenerational consistency between the smartwatch IMUs. This provides a measure of confidence to pool and compare studies using IMU data recorded from different versions of the smartwatch.

The sensor signal quality experiment had several strengths. In contrast to earlier studies validating consumer sensor proprietary ‘black-box’ energy expenditure, heart rate, step count measures, and joint estimates, our study was unique in measuring the accuracy of the straightforward inertial movement measurements (i.e. acceleration and angular velocity) of the sensors. We were able to do this by developing custom software to bypass the consumer smartwatch closed system barriers to export raw acceleration and angular velocity data. Using our custom extraction and transmission of the smartwatch IMU data, the data could easily be integrated with wider systems for unique research and clinical applications outside of the laboratory. Studies [50-52] also developed custom software to export Apple Watch data but did not assess the IMU accuracy. Moreover, our validation of 4 individual Xsens MTw sensor accuracy, as opposed to the 17-sensor Xsens MVN BIOMECH full-body suit in earlier studies, is also noteworthy. We posit that the full body suit is less practical for long-term continuous ‘in the wild’ behaviour monitoring of in-patients as it is higher cost, more challenging to operate and calibrate, obtrusive for the user for long wear times (requires 17 sensors attached to various positions on the body), and requires mobile participants for the walking calibration. We evaluated whether the position of the sensor in the sensor stack (i.e. distance to the wrist) affected the captured movement estimates and found that it was not a significant confound.

Limitations of our sensor accuracy study include the fact that the movement task duration (∼5 minutes) may not have sufficiently captured drift over longer time-periods and the controlled task of the upper limb may not have fully represented the range of natural human movements. Importantly, we note that some differences between optical motion tracking and Xsens could be explained by greater inertial sensor jitter and latency and horizontal position (XY) drift during stationary periods or exaggerated noise from marker obstruction through the differentiation when deriving acceleration and angular velocity from optical motion tracking position data [53].

In summary, our sensor quality study results demonstrate that: 1. sensors with both accelerometers and gyroscopes (Xsens and Apple Watches) perform better than just accelerometers (Axivity); 2. accuracy for acceleration, but not angular velocity, is higher for research-grade sensors that employ SDI technology with magnetometry (Xsens), compared to non-SDI non-magnetometer sensors (Apple Watches); 3. Apple Watch Series 3 and Series 5 demonstrated intergenerational inertial sensing consistency i.e. were practically identical, suggesting. This gave us confidences that, with relatively few drawbacks, consumer-grade smartwatches can be objectively used within clinical- and research-grade setting.

### 4.2. Sensor Acceptability Study

Similarly, our acceptability results show that subjectively consumer-grade smartwatches are suitable to be used within clinical- and research-grade environments. The sensor acceptability study demonstrated high approval ratings from hospital patients and healthcare professionals for use of wearable motion sensors for continuous motion tracking. We found that in-patients were generally neutral towards or had nothings comment about the sensors in open responses; and strongly agreed with closed-ended statements that the sensors were simple to use, comfortable, unobtrusive and did not interfere with daily activities. At the same time, a small number of participants did raise worries with regards to discomfort, bulky sensor size, data privacy, and damage and loss. These concerns were similarly expressed across earlier wearable sensor usability studies [13,23]. For example, Tran and authors (2019) found that patient’s data privacy concerns including hacking of data and devices, spying on patients, and using and selling patient data without consent. Our findings highlighted that patients were considerably influenced by superficial characteristics related to sensor design and appearance, such as sensor colour schemes. This mirrors earlier findings, such as [21] which reported positive user opinions with regards to ‘colourful’, ‘beautiful’, ‘lightweight’ designs and ‘ease of use’ of evaluated smartwatches and [23] which recorded ‘lack of attractive features’ as the top concern for wearable devices. Acceptability results from healthcare professionals also revealed conflicting views regarding wearable sensor motion tracking across different medical specialities, such as the perceived opportunity of wearable movement sensors and artificial intelligence in healthcare applications. This highlights that different members of multidisciplinary teams have different experiences and expectations of technologies such as motion trackers, which need to be addressed when introducing such systems into clinical environments.

The device feasibility study had several strengths. Decision to adopt or use a new technology frequently involves a shared agreement between both the patients and healthcare professionals. Our study looked at both perspectives through the questionnaires. Our combined methods design, using open-ended and closed-ended questions, allowed broad insights into perceptions of the use of wearable sensors for continuous monitoring in a hospital setting. Comparable to earlier smartwatch usability studies [21], we used a Likert scale evaluation to enable us to gauge degrees of opinion. Furthermore, the findings captured a diverse range of views from a large sample of in-patients (n=44) with wide-ranging demographics and multiple comorbidities. This offers an advantage over earlier studies such as [21] which only captured views from 7 healthy subjects in a community setting; [22,23] which collected data from larger samples (n=388 and n=2058 respectively), but only recruited healthy subjects online who reported some or no experience using the wearable wristwatches.

The feasibility study was limited by not using a validated device usability questionnaire, such as System Usability Scale (SUS)[28], as we wanted to explore a broader set of questions (as subjects were not using the wearables as such but wearing them) while keeping questions brief to ensure completion and compliance rates. Unlike study [21], we did not include the evaluation of satisfaction with the smartwatch user interface or battery as part of the study and prevented user on-screen interactions. Given that we wanted to evaluate the inertial sensor primarily for recording movement for healthcare professional in-patient monitoring, we did not choose to assess in-patient views of other smartwatch features and functionalities (e.g. networking and applications). In future research, we plan to develop and assess the acceptability of custom device movement feedback visualisations. Our smaller sample size of healthcare professionals (n=15) may not be representative of the broader population and the sample was unbalanced for different medical specialities and experience. Views on the usability of wearables were based upon recording durations (1.5-24hour) which is shorter than typical in-patient stays of several days to weeks. Our study also gained opinions only from those of patients in neurology and stroke wards, which may not be representative of other clinical settings and care homes. The study only collected views on Apple Watch wearables, which may not generalise to other wearable inertial devices. However, this allowed us to incorporate brand-related influences (e.g. brand loyalty and attitudes) that play a role in end-user adoption and to address a gap in the literature for perceptions related to use consumer devices for continuous monitoring in hospital settings [54]. We believe that there is, in principle, no technological barriers in allowing other smartwatch platforms with appropriate programmable interfaces and high-accuracy IMUs to be developed and look forward to this rapidly developing consumer electronics domain developing common interoperability standards for measurement, collecting and deploying of healthcare data and applications.

In summary, our sensor acceptability study showed that 1) hospital patients wearing motion tracking smartwatches for 1.5-24 hours are positive about their use; 2) healthcare professionals involved in clinical monitoring also embraced wearable IMU technology but concerns that need to be addressed are data privacy, compliance, sensor loss, specificity and discomfort.

## 5. Conclusions

These results suggest that for continuous long-term behavioural monitoring of in-patients, consumer smartwatches (such as Apple Watch) can offer reliable inertial tracking. Albeit more so for measures relying on angular velocity, than linear acceleration for Apple smartwatches. The implication of this on clinical application, for example, to measure the proportion of time lying in bed, as opposed to ambulating, or to estimate physical disability, needs to be ascertained by further studies. Our feasibility results provide reassurance that consumer smartwatch motion tracking is generally acceptable for patients and staff in hospitals, where we can now proceed with deploying these consumer-grade technologies to easily collect and monitor natural behaviour on a daily-basis from in-patient and care home residents. This may pay the wave for improved care, patient safety, and novel data-driven solutions enabled by the availability of low-cost, high-accuracy natural behavioural data streams (e.g. [55]) that can be collected in a low-cost, accurate, continuous and socially distanced manner. Our results show that consumer-grade smartwatch use effectively provides researchers and healthcare technology developers with an accurate and acceptable platform enabling to 24/7 watch over a patient.

## Data Availability

The raw sensor data traces can be made available upon request to the corresponding authors. The survey answer numbers have been published here in full.

## Supplementary Materials

The following are available online at www.mdpi.com/xxx/A1, Table A1: Movement Sequence Protocol, Table A2: In-patient characteristics (n =44), Table A3: Healthcare professional characteristics (n =15), and Figure A1.

## Author Contributions

For research articles with several authors, a short paragraph specifying their individual contributions must be provided. The following statements should be used “Conceptualization, PB and AAF; methodology, AAF, PB, SW and CA; software, AAF and CA; validation, AAF, PB, SW and CA; formal analysis, AAF, CA and SW; experimental data collection, CA, SW, and JS; resources, PB and AAF; data curation, SW, CA, and JS; writing—original draft preparation, SW and CA; writing—review and editing AAF, PB, SW and CA; visualisation, SW and CA; supervision, PB and AAF; project administration, PB and AAF; funding acquisition, PB and AAF.

## Acknowledgements

We would like to thank Dr Balasundaram Kadirvelu for his advice and his help in setting up the experiments. We also would like to thank all the participants for taking part in our two studies.

### Appendix A

**Table A1.** Movement Sequence Protocol

| Action | TPA<sup>2</sup> | BPA<sup>1</sup> | REPS<sup>3</sup> | TOT<sup>4</sup> |
| --- | --- | --- | --- | --- |
| **Handclap** |  |  |  |  |
| • shoulder 45 degrees, elbow 45 degrees | 0.5 | 1 | 5 | 2.5 |
| **Horizontal Arm Movement (Both arms)** |  |  |  |  |
| • hand to contralateral shoulder, elbow 0-90 degrees, forearm pronated, shoulder at 90 degrees | 2 | 4 | 30 | 60 |
| **Vertical Arm Movement (Both arms)** |  |  |  |  |
| • Shoulder 0-90 degrees vertical abduction-adduction, elbow 0 degrees, forearm pronated | 2 | 4 | 30 | 60 |
| • Shoulder 0-90 degrees vertical flexion-extension, elbow 0 degrees, forearm pronated | 2 | 4 | 30 | 60 |
| **Rotational Arm Movement** |  |  |  |  |
| • Pronation-supination, shoulder 90 degrees vertical flexion, elbow 0 degrees | 1 | 2 | 30 | 30 |
| • Pronation-Supination, shoulder 90 degrees vertical flexion, elbow 90 degrees vertical | 1 | 2 | 30 | 30 |
| • Pronation-Supination, shoulder 90 degrees vertical flexion, elbow 90 degrees horizontal | 1 | 2 | 30 | 30 |
| **Composite Cross-Body Movement** |  |  |  |  |
| • Hand from contralateral knee to ipsilateral ear, back bent | 1 | 2 | 30 | 30 |
| **Handclap** |  |  |  |  |
| • shoulder 45 degrees, elbow 45 degrees | 0.5 | 1 | 5 | 2.5 |
| Total time (Not including rest periods between each action) |  | 305 seconds |  |  |

Each movement task consists of the movement task description, <sup>1</sup> number of metronome beats per task (BPA),
<sup>2</sup> The Time Per Action (TPA) (calculated based on 120 Beat Per Minute frequency and length of action cycle),
<sup>3</sup> number of movement task repetitions (REP), and <sup>4</sup> total times of the movement task (TOT).

**Table A2.** In-patient characteristics (n =44)

| In-Patient Characteristics | n (weights) |
| --- | --- |
| Age (years)— *Avg (IQR)* | 64 (24–92) |
| Female sex — *n (%)* | 22 (50) |
| Male sex — *n (%)* | 22 (50) |
| Asthma — *n (%)* | 4 (9) |
| Chronic obstructive pulmonary disease — *n (%)* | 3 (7) |
| Other respiratory diseases — *n (%)* | 2 (5) |
| Diabetes — *n (%)* | 4 (9) |
| Thyroid disorders — *n (%)* | 3 (7) |
| High blood pressure — *n (%)* | 29 (66) |
| Dyslipidemia — *n (%)* | 13 (30) |
| Other cardiac or vascular diseases — *n (%)* | 5 (11) |
| Chronic kidney diseases — *n (%)* | 2 (5) |
| Rheumatologic conditions — *n (%)* | 6 (13) |
| Digestive conditions — *n (%)* | 10 (23) |
| Neurological conditions — *n (%)* | 4 (9) |
| Cancer (including blood cancer) — *n (%)* | 2 (5) |
| Depression — *n (%)* | 2 (5) |
| Ischemic stroke — *n (%)* | 29 (66) |
| Hemorrhagic stroke — *n (%)* | 5 (11) |
| Transient Ischemic Attack — *n (%)* | 10 (23) |
| Stroke mimic (Stroke symptoms but non-stroke) — *n (%)* | 10 (23) |

Avg = Average; IQR = Interquartile range; n = number

**Table A3.** Healthcare professional characteristics (n =15)

| Characteristics | Doctor (n=5) | Nurses (n=4) | HCA (n=3) | Therapist (n=3) |
| --- | --- | --- | --- | --- |
| Age (years)— *Avg*<br>(*IQR*) | 28 (27–32) | 40 (31–58) | 48 (30–55) | 33 (28–45) |
| Female sex — *n* (%) | 2 (40%) | 2 (50%) | 3 (100%) | 3 (100%) |
| Male sex — *n* (%) | 3 (60%) | 2 (50%) | 0 (0%) | 0 (0%) |
| Educational level (%) | Medical degree<br>(60%), MSc (40%) | Degree level<br>(100%) | Degree level<br>(33%), NVQ3<br>(33%), N/A (33%) | Degree (66%),<br>N/A (33%) |
| Clinical speciality or<br>training level | 1 Stroke SHO, 1 CT<br>medicine, 1<br>Geriatrics ST3, 1<br>GPS 2, 1 SPR | Student Nurse<br>(25%), B5<br>Nurse (75%) | HCA level 2<br>(33%), N/A (66%) | 1 Rehab assistant,<br>1 B5<br>Occupational<br>therapist, 1 B6<br>Physiotherapist |
| Previous use of<br>e-health or m-health<br>technology | Yes (40%), No (60%) | Yes (25%), No<br>(75%) | No (100%) | Yes (33%), No<br>(66%) |

*Avg* = Average; *IQR* = Interquartile range, *n* = number; SPR = Specialist Registrar; SHO = Senior House Officer; ST3 = Specialty Training 3; CT = Core Training; NVQ = National Vocational Qualifications; MSc = Master of Science; GPS = General Practice Specialty

**Figure A1.**
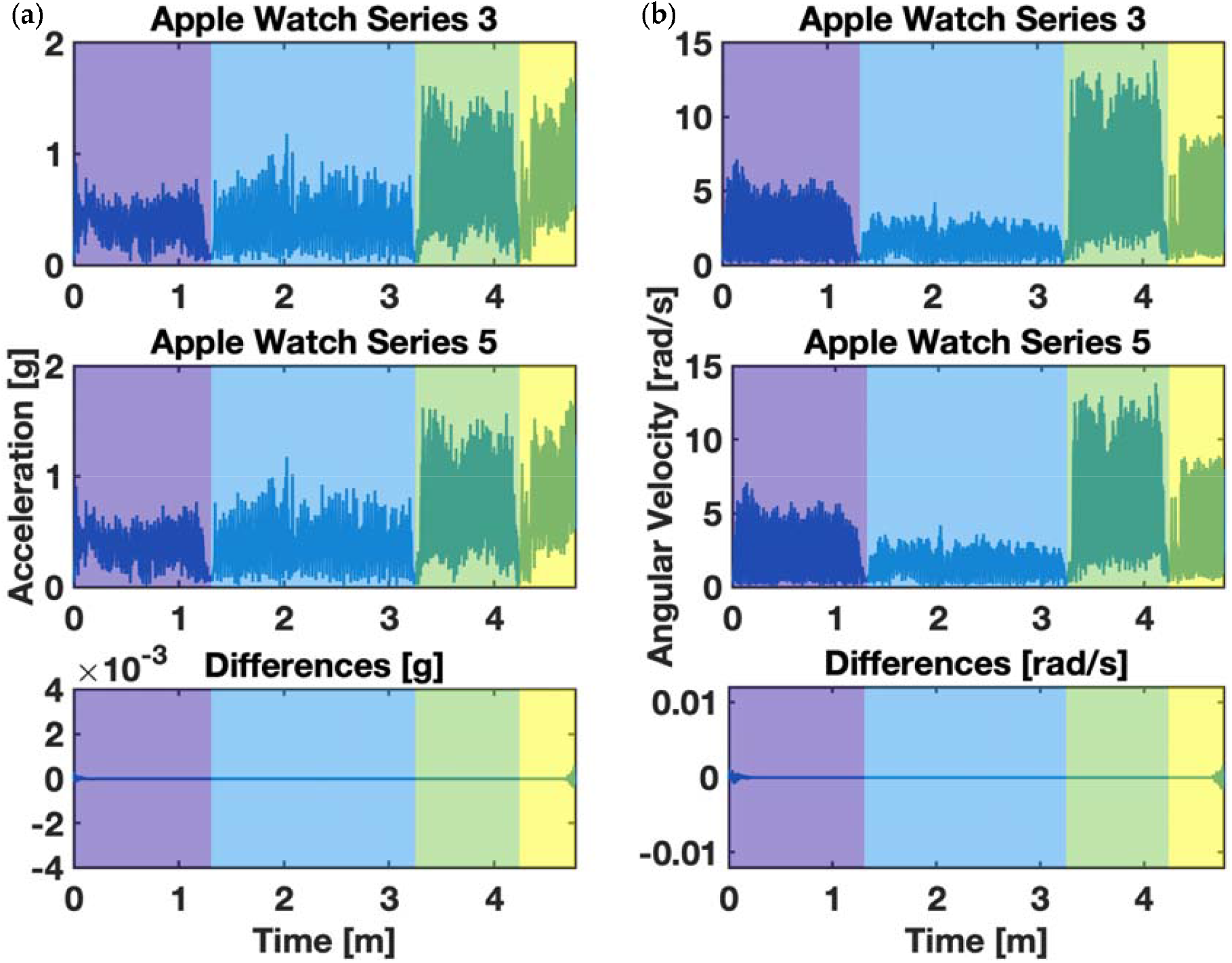
depicts recorded IMU signal from Apple Watch Series 5 (top of stack) and Apple Watch Series 3 (bottom of the stack) where (a) are accelerations and their differences and (b) are angular velocities and their differences. The coloured area indicates different behavioural execution as depicted in Figure 3 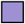 i.e. is 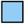 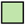 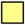 horizontal arm movements, is vertical arm movements, is rotational arm movements, and is composite cross-body movements.

## References

1. Salter, K., Campbell, N., Richardson, M., Mehta, S., Jutai, J., Zettler, L., Moses, M.B.A., McClure, A., Mays, R., Foley, N., et al. EBRSR [Evidence-Based Review of Stroke Rehabilitation] 20 Outcome Measures in Stroke Rehabilitation.

2. Johansson, D., Malmgren, K., Alt Murphy, M. Wearable sensors for clinical applications in epilepsy, Parkinson’s disease, and stroke: a mixed-methods systematic review. 2018; 10.1007/s00415-018-8786-y.

3. Kim, C.M., Eng, J.J. Magnitude and pattern of 3D kinematic and kinetic gait profiles in persons with stroke: relationship to walking speed. Gait Posture 2004, 20, 140–146, doi:10.1016/j.gaitpost.2003.07.002.

4. Cuesta-Vargas, A.I., Galan-Mercant, A., Williams, J.M. The use of inertial sensors system for human motion analysis. Phys Ther Rev 2010, 15, 462–473, doi:10.1179/1743288X11Y.0000000006.

5. Kavanagh, J.J., Menz, H.B. Accelerometry: a technique for quantifying movement patterns during walking. Gait Posture 2008, 28, 1–15, doi:10.1016/j.gaitpost.2007.10.010.

6. Patel, S., Park, H., Bonato, P., Chan, L., Rodgers, M. A review of wearable sensors and systems with application in rehabilitation. Journal of NeuroEngineering and Rehabilitation 2012, 9, 21, doi:10.1186/1743-0003-9-21.

7. Maetzler, W., Domingos, J., Srulijes, K., Ferreira, J.J., Bloem, B.R. Quantitative wearable sensors for objective assessment of Parkinson’s disease. Mov Disord 2013, 28, 1628–1637, doi:10.1002/mds.25628.

8. Sim, N., Gavriel, C., Abbott, W.W., Faisal, A.A. The head mouse@ Head gaze estimation “In-the-Wild” with low-cost inertial sensors for BMI use. 2013 6th International IEEE/EMBS Conference on Neural Engineering (NER) 2013, 735–738.

9. Gavriel, C., Faisal, A.A. Wireless kinematic body sensor network for low-cost neurotechnology applications “in-the-wild”. In Proceedings of 2013 6th International IEEE/EMBS Conference on Neural Engineering (NER), 6-8 Nov. 2013; pp. 1279–1282.

10. Lopez-Nava, I.H., Angelica, M.M. Wearable Inertial Sensors for Human Motion Analysis: A review. Institute of Electrical and Electronics Engineers Inc.: 2016; Vol. PP.

11. Reeder, B., David, A. Health at hand: A systematic review of smart watch uses for health and wellness. J Biomed Inform 2016, 63, 269–276, doi:10.1016/j.jbi.2016.09.001.

12. King, C.E., Sarrafzadeh, M. A Survey of Smartwatches in Remote Health Monitoring. Journal of Healthcare Informatics Research 2018, 2, 1–24, doi:10.1007/s41666-017-0012-7.

13. Piwek, L., Ellis, D.A., Andrews, S., Joinson, A. The Rise of Consumer Health Wearables: Promises and Barriers. PLoS Med 2016, 13, e1001953, doi:10.1371/journal.pmed.1001953.

14. Wallen, M.P., Gomersall, S.R., Keating, S.E., Wisloff, U., Coombes, J.S. Accuracy of Heart Rate Watches: Implications for Weight Management. PLoS One 2016, 11, e0154420, doi:10.1371/journal.pone.0154420.

15. Xie, J., Wen, D., Liang, L., Jia, Y., Gao, L., Lei, J. Evaluating the Validity of Current Mainstream Wearable Devices in Fitness Tracking Under Various Physical Activities: Comparative Study. JMIR Mhealth Uhealth 2018, 6, e94, doi:10.2196/mhealth.9754.

16. Evenson, K.R., Goto, M.M., Furberg, R.D. Systematic review of the validity and reliability of consumer-wearable activity trackers. Int J Behav Nutr Phys Act 2015, 12, 159, doi:10.1186/s12966-015-0314-1.

17. Dooley, E.E., Golaszewski, N.M., Bartholomew, J.B. Estimating Accuracy at Exercise Intensities: A Comparative Study of Self-Monitoring Heart Rate and Physical Activity Wearable Devices. JMIR Mhealth Uhealth 2017, 5, e34, doi:10.2196/mhealth.7043.

18. An, H.S., Jones, G.C., Kang, S.K., Welk, G.J., Lee, J.M. How valid are wearable physical activity trackers for measuring steps? Eur J Sport Sci 2017, 17, 360–368, doi:10.1080/17461391.2016.1255261.

19. Kooiman, T.J., Dontje, M.L., Sprenger, S.R., Krijnen, W.P., van der Schans, C.P., de Groot, M. Reliability and validity of ten consumer activity trackers. BMC Sports Sci Med Rehabil 2015, 7, 24, doi:10.1186/s13102-015-0018-5.

20. Shcherbina, A., Mattsson, C.M., Waggott, D., Salisbury, H., Christle, J.W., Hastie, T., Wheeler, M.T., Ashley, E.A. Accuracy in Wrist-Worn, Sensor-Based Measurements of Heart Rate and Energy Expenditure in a Diverse Cohort. J Pers Med 2017, 7, pdoi:10.3390/jpm7020003.

21. Kaewkannate, K., Kim, S. A comparison of wearable fitness devices. BMC Public Health 2016, 16, 433, doi:10.1186/s12889-016-3059-0.

22. Liang, J., Xian, D., Liu, X., Fu, J., Zhang, X., Tang, B., Lei, J. Usability Study of Mainstream Wearable Fitness Devices: Feature Analysis and System Usability Scale Evaluation. JMIR Mhealth Uhealth 2018, 6, e11066, doi:10.2196/11066.

23. Wen, D., Zhang, X., Lei, J. Consumers’ perceived attitudes to wearable devices in health monitoring in China: A survey study. Comput Methods Programs Biomed 2017, 140, 131–137, doi:10.1016/j.cmpb.2016.12.009.

24. Tran, V.-T., Riveros, C., Ravaud, P. Patients’ views of wearable devices and AI in healthcare: findings from the ComPaRe e-cohort. npj Digital Medicine 2019, 2, 53–53, doi:10.1038/s41746-019-0132-y.

25. Hsiao, K.-L., Chen, C.-C. What drives smartwatch purchase intention? Perspectives from hardware, software, design, and value. Telematics and Informatics 2018, 35, 103–113, doi:10.1016/j.tele.2017.10.002.

26. Market share of smartwatch unit shipments worldwide from the 2Q’14 to 1Q ‘20*, by vendor. Availabe online: https://www.statista.com/statistics/524830/global-smartwatch-vendors-market-share/ (xaccessed on 22 June).

27. Zhao, Y., Heida, T., Van Wegen, E.E.H., Bloem, B.R., Van Wezel, R.J.A. E-health support in people with Parkinson’s disease with smart glasses: A survey of user requirements and expectations in the Netherlands. Journal of Parkinson’s Disease 2015, 10.3233/JPD-150568, doi:10.3233/JPD-150568.

28. Brooke, J. SUS: a retrospective. J. Usability Studies 2013, 8, 29–40.

29. Zhang, J.-T., Novak, A.C., Brouwer, B., Li, Q. Concurrent validation of Xsens MVN measurement of lower limb joint angular kinematics. Physiological Measurement 2013, 34, N63–N69, doi:10.1088/0967-3334/34/8/n63.

30. Thies, S.B., Tresadern, P., Kenney, L., Howard, D., Goulermas, J.Y., Smith, C., Rigby, J. Comparison of linear accelerations from three measurement systems during “reach & grasp”. Med Eng Phys 2007, 29, 967–972, doi:10.1016/j.medengphy.2006.10.012.

31. Cloete, T., Scheffer, C. Benchmarking of a full-body inertial motion capture system for clinical gait analysis. 2008 30th Annual International Conference of the IEEE Engineering in Medicine and Biology Society 2008, 4579–4582.

32. Robert-Lachaine, X., Mecheri, H., Larue, C., Plamondon, A. Validation of inertial measurement units with an optoelectronic system for whole-body motion analysis. Medical & Biological Engineering & Computing 2017, 55, 609–619, doi:10.1007/s11517-016-1537-2.

33. Teufl, W., Lorenz, M., Miezal, M., Taetz, B., Frohlich, M., Bleser, G. Towards Inertial Sensor Based Mobile Gait Analysis: Event-Detection and Spatio-Temporal Parameters. Sensors (Basel) 2018, 19, doi:10.3390/s19010038.

34. Teufl, W., Miezal, M., Taetz, B., Frohlich, M., Bleser, G. Validity, Test-Retest Reliability and Long-Term Stability of Magnetometer Free Inertial Sensor Based 3D Joint Kinematics. Sensors (Basel) 2018, 18, doi:10.3390/s18071980.

35. Karatsidis, A., Jung, M., Schepers, H.M., Bellusci, G., de Zee, M., Veltink, P.H., Andersen, M.S. Musculoskeletal model-based inverse dynamic analysis under ambulatory conditions using inertial motion capture. Med Eng Phys 2019, 65, 68–77, doi:10.1016/j.medengphy.2018.12.021.

36. Del Din, S., Godfrey, A., Rochester, L. Validation of an Accelerometer to Quantify a Comprehensive Battery of Gait Characteristics in Healthy Older Adults and Parkinson’s Disease: Toward Clinical and at Home Use. IEEE J Biomed Health Inform 2016, 20, 838–847, doi:10.1109/JBHI.2015.2419317.

37. Godfrey, A., Del Din, S., Barry, G., Mathers, J.C., Rochester, L. Instrumenting gait with an accelerometer: A system and algorithm examination. Medical Engineering & Physics 2015, 37, 400–407, doi:https://doi.org/10.1016/j.medengphy.2015.02.003.

38. Doherty, A., Jackson, D., Hammerla, N., Plotz, T., Olivier, P., Granat, M.H., White, T., van Hees, V.T., Trenell, M.I., Owen, C.G., et al. Large Scale Population Assessment of Physical Activity Using Wrist Worn Accelerometers: The UK Biobank Study. PLoS One 2017, 12, e0169649, doi:10.1371/journal.pone.0169649.

39. Carse, B., Meadows, B., Bowers, R., Rowe, P. Affordable clinical gait analysis: An assessment of the marker tracking accuracy of a new low-cost optical 3D motion analysis system. Physiotherapy 2013, 99, 347–351, doi:10.1016/j.physio.2013.03.001.

40. Ehara, Y., Fujimoto, H., Miyazaki, S., Mochimaru, M., Tanaka, S., Yamamoto, S. Comparison of the performance of 3D camera systems II. Gait and Posture 1997, 5, 251–255, doi:10.1016/S0966-6362(96)01093-4.

41. Maciejewski, M., Piszczek, M., Pomianek, M. Testing the SteamVR trackers operation correctness with the OptiTrack system; SPIE: 2018; Vol. 10830.

42. Mortazavi, B., Nemati, E., Wall, K.V., Flores-Rodriguez, H.G., Cai, J.Y.J., Lucier, J., Naeim, A., Sarrafzadeh, M. Can smartwatches replace smartphones for posture tracking? Sensors (Switzerland) 2015, 15, 26783–26800, doi:10.3390/s151026783.

43. Cuesta-Vargas, A.I., Galán-Mercant, A., Williams, J.M. The use of inertial sensors system for human motion analysis. Taylor and Francis Ltd.: 2010; Vol. 15, pp 462–473.

44. Roetenberg, D., Slycke, P.J., Veltink, P.H. Ambulatory Position and Orientation Tracking Fusing Magnetic and Inertial Sensing. IEEE Transactions on Biomedical Engineering 2007, 54, 883–890, doi:10.1109/TBME.2006.889184.

45. O’Donovan, K.J., Kamnik, R., O’Keeffe, D.T., Lyons, G.M. An inertial and magnetic sensor based technique for joint angle measurement. J Biomech 2007, 40, 2604–2611, doi:10.1016/j.jbiomech.2006.12.010.

46. Zhang, M., Hol, J.D., Slot, L., Luinge, H. Second order nonlinear uncertainty modeling in strapdown integration using MEMS IMUs; 2011; pp. 1–7.

47. Bortz, J.E. A New Mathematical Formulation for Strapdown Inertial Navigation. IEEE Transactions on Aerospace and Electronic Systems 1971, AES-7, 61–66, doi:10.1109/TAES.1971.310252.

48. Paulich, M., Schepers, M., Rudigkeit, N., Bellusci, G. Xsens MTw Awinda: Miniature Wireless Inertial-Magnetic Motion Tracker for Highly Accurate 3D Kinematic Applications; 2018; 10.13140/RG.2.2.23576.49929.

49. Bergamini, E., Ligorio, G., Summa, A., Vannozzi, G., Cappozzo, A., Sabatini, A.M. Estimating orientation using magnetic and inertial sensors and different sensor fusion approaches: accuracy assessment in manual and locomotion tasks. Sensors (Basel) 2014, 14, 18625–18649, doi:10.3390/s141018625.

50. Walch, O., Huang, Y., Forger, D., Goldstein, C. Sleep stage prediction with raw acceleration and photoplethysmography heart rate data derived from a consumer wearable device. Sleep 2019, 42, pdoi:10.1093/sleep/zsz180.

51. Amroun, H., Temkit, M., Ammi, M. DNN-Based Approach for Recognition of Human Activity Raw Data in Non-Controlled Environment. In Proceedings of 2017 IEEE International Conference on AI & Mobile Services (AIMS); pp. 121–124.

52. Kwon, M.C., Park, G., Choi, S. Smartwatch User Interface Implementation Using CNN-Based Gesture Pattern Recognition. Sensors (Basel) 2018, 18, doi:10.3390/s18092997.

53. Esser, P., Dawes, H., Collett, J., Howells, K. IMU: inertial sensing of vertical CoM movement. J Biomech 2009, 42, 1578–1581, doi:10.1016/j.jbiomech.2009.03.049.

54. Chuah, S.H.-W., Rauschnabel, P.A., Krey, N., Nguyen, B., Ramayah, T., Lade, S. Wearable technologies: The role of usefulness and visibility in smartwatch adoption. Computers in Human Behavior 2016, 65, 276–284, doi:10.1016/j.chb.2016.07.047.

55. Xiloyannis, M., Gavriel, C., Thomik, A.A.C., Faisal, A.A. Gaussian Process Autoregression for Simultaneous Proportional Multi-Modal Prosthetic Control With Natural Hand Kinematics. IEEE Transactions on Neural Systems and Rehabilitation Engineering 2017, 25, 1785–1801, doi:10.1109/TNSRE.2017.2699598.

